# Wastewater surveillance of human influenza, metapneumovirus, parainfluenza, respiratory syncytial virus (RSV), rhinovirus, and seasonal coronaviruses during the COVID-19 pandemic

**DOI:** 10.1101/2022.09.22.22280218

**Authors:** Alexandria B. Boehm, Bridgette Hughes, Dorothea Duong, Vikram Chan-Herur, Anna Buchman, Marlene K. Wolfe, Bradley J. White

## Abstract

**Background:** Respiratory disease is a major cause of morbidity and mortality; however, current surveillance for circulating respiratory viruses is passive and biased. Seasonal circulation of respiratory viruses changed dramatically during the COVID-19 pandemic. More active methods for understanding respiratory disease dynamics are needed to better inform public health response and to guide clinical decision making. Wastewater-based epidemiology has been used to understand COVID-19, influenza A, and RSV infection rates at a community level, but has not been used to investigate other respiratory viruses.

**Methods:** We measured concentrations of influenza A and B, RSV A and B, human parainfluenza (1-4), rhinovirus, seasonal human coronaviruses, and human metapneumovirus RNA in wastewater solids three times per week for 17 months spanning the COVID-19 pandemic at a wastewater treatment plant in California, USA. Novel probe-based assays were developed and validated for non-influenza viral targets. We compared viral concentrations to positivity rates for viral infections from clinical specimens submitted to sentinel laboratories.

**Findings:** We detected RNA from all target viruses in wastewater solids. Human rhinovirus and seasonal coronaviruses were found at highest concentrations. Concentrations of viruses correlated significantly and positively with positivity rates of associated viral diseases from sentinel laboratories. Measurements from wastewater indicated limited circulation of RSV A and influenza B, and human coronavirus OC43 dominated the seasonal human coronavirus infections while human parainfluenza 1 and 4A dominated among parainfluenza infections.

**Interpretation:** Wastewater-based epidemiology can be used to obtain information on circulation of respiratory viruses at a community level without the need to test many individuals because a single sample of wastewater represents the entire contributing community. Results from wastewater can be available within 24 hours of sample collection, allowing real time information to inform public health response, clinical decision making, and individual behavior modifications.

## Introduction

Acute respiratory illnesses (ARI) are defined as symptomatic infections of the lower or upper respiratory tract, and may also result in systemic symptoms and secondary infections. They are consistently the top cause of illness and mortality in children under 5 globally after the neonatal period, and represent a large burden of infectious disease^1^. ARI etiology can be bacterial, fungal, or viral; lower respiratory infections most commonly lead to severe conditions like pneumonia and bronchiolitis, while upper respiratory infections are largely self limiting but can result in secondary infections. Viral ARI are most commonly caused by SARS-CoV-2, seasonal human coronavirus, human parainfluenza, human metapneumovirus, influenza, respiratory syncytial virus, rhinovirus, adenoviruses, human bocaviruses, and non-rhinovirus enteroviruses^2^. In the United States, viral ARI causes nearly 400,000 child hospitalizations at a cost of $1 billion annually^3^.

Respiratory disease surveillance in the United States is passive and draws from many sources, often relying on institutions to identify specific diseases through key words in clinical records, testing of clinical specimens, and searching for keywords on death certificates. The passive respiratory disease surveillance system is voluntary and biased towards identifying infections in individuals with co-morbidities and/or where symptoms are severe and patients require clinical care^4^. Since many ARIs are self-limiting, diagnostic testing of most individuals with respiratory illness is uncommon. Lack of robust, unbiased data on disease occurrence and resultant lack of knowledge on occurrence and trends in circulating respiratory diseases limits public health response, institutional awareness that can guide clinical decision making regarding testing and treatment, and efforts to understand disease epidemiology including disease interference and co-infection^5^.

Prior to the COVID-19 pandemic, viral respiratory infections largely circulated seasonally with predictable dynamics^2^. However, seasonal occurrence of respiratory viruses has changed dramatically during the COVID-19 pandemic^6^, presumably due to both imposed and voluntary behavior changes in response to the pandemic, such as masking and stay-at-home orders^7^. It is uncertain if and when respiratory disease dynamics will return to those observed pre-pandemic. More active approaches to disease surveillance are needed to better understand and respond to atypical respiratory disease dynamics.

Wastewater-based epidemiology (WBE) uses the information from wastewater to gain insight into infectious disease occurrence in contributing communities. Wastewater represents a composite biological sample containing bodily excretions including urine, feces, sputum, and mucus. Excretions of infected individuals can contain markers of infectious disease (including live and dead organisms, proteins and nucleic acids) and methods have been developed to detect and quantify these in the wastewater matrix. Benefits of WBE are that it does not require that infected individuals receive medical care or testing or even experience symptoms to be represented in the community-level data source.

The utility of WBE has been demonstrated for understanding community circulation of poliovirus^8^, *Salmonella*^9^, hepatitis A^10^, and enteroviruses^11^, but its use has not widely been a part of disease surveillance outside of polio in endemic regions. The application of WBE to respiratory diseases is relatively new and was prompted by the COVID-19 pandemic, during which it was widely shown that concentrations of SARS-CoV-2 RNA in wastewater correlate to reported COVID-19 incidence rates during periods of widely accessible clinical testing^12^. Thereafter, Hughes et al.^13^ and Wolfe et al.^14^ showed RSV and influenza A RNA concentrations, respectively, in wastewater solids were associated with disease occurrence in associated sewersheds.

Here we test the utility of WBE for a suite of viral respiratory diseases. We developed and validated novel hydrolysis probe-based RT-PCR assays that target respiratory viral genomes and then applied the assays to wastewater solids collected three times per week at a wastewater treatment plant over 17 months during the COVID-19 pandemic. We measured concentrations of respiratory syncytial virus (RSV) A and B, influenza A and B, metapneumovirus, parainfluenza (1-4), seasonal human coronaviruses (229E, OC43, NL63, and HKU-1), and rhinovirus RNA. We chose to focus on these respiratory viruses as passive clinical surveillance occurs for these diseases in the region where the sampling was conducted allowing us to compare wastewater to clinical surveillance data. Additionally, these viruses represent important and common viral etiologies of respiratory infections^15^.

## Methods

### RT-PCR assays

We used published hydrolysis probe-based RT-PCR assays for Influenza A (IAV) and influenza B (IBV)^16^. We designed novel RT-PCR primers and internal hydrolysis probes for rhinovirus (hereafter, “HRV” targeting rhinovirus A, B and C together); human parainfluenza (HPIV) 1, 2, 3, 4A, 4B; human coronavirus (HCoV) 229E, NL63, OC43, HKU-1; human metapneumovirus (HMPV); and Respiratory Syncytial Virus (RSV) A and B. To develop the assays, viral genome sequences were downloaded from NCBI between between February and June 2022 and aligned to identify conserved regions. Assays were developed *in silico* using Primer3Plus (https://primer3plus.com/). Parameters used in assay development (e.g. sequence length, GC content, and melt temperatures) are provided in Table S1. Primers and probes were then screened for specificity *in silico*, and *in vitro* against virus panels, intact respiratory viruses, synthetic viral genomic RNA, or cDNA sequences (see Supplementary Material (SM) including Table S2).

### Wastewater analyses

A wastewater treatment plant that serves 75% (1,500,000 people) of Santa Clara County, California (San José-Santa Clara Regional Wastewater Facility, RWF) was included in the study. Further descriptions of RWF can be found in elsewhere^12^.

Samples were collected daily for a COVID-19 WBE effort starting in November 2020^12^, and a subset of those samples (three samples per week, 216 samples total) are used in this study. The samples were chosen to span a 17 month period (2/2/21 - 6/21/22, month/day/year format) that included implementation and easing of indoor mask mandates, changes in COVID-19 vaccine availability, public health promotion of hand hygiene and mask wearing, and periods of both high and low COVID-19 incidence.

Fifty mL of settled solids were collected using sterile technique in clean bottles from the primary clarifier. Twenty-four hour composite samples were collected by combining grab samples from the sludge line every six hours^12^. Samples were stored at 4°C, transported to the lab, and processed within six hours. Solids were then dewatered^12^ and frozen at −80°C for 4 - 60 weeks. Samples were thawed overnight and then RNA was obtained from the dewatered solids following previously published protocols^12^ (see SM). RNA was obtained from 10 replicate sample aliquots. Each replicate RNA extract from each sample was subsequently processed immediately to measure viral RNA concentrations using digital RT-PCR with multiplexed assays for IAV, IBV, HMPV, total HPIV (HPIV 1, HPIV 2, HPIV 3, HPIV 4A, and HPIV 4B), total HCoV (HCoV 229E, HCoV NL63, HCoV OC43, HCoV HKU-1), RSV A, RSV B, HRV, and pepper mild mottle virus (PMMoV) (see SI). PMMoV is highly abundant in human stool and wastewater globally^17^ and is used as an internal recovery and fecal strength control^18^. Nine samples spanning the duration of the time series were selected to measure each HCoV and HPIV individually (Table S3). Each 96-well PCR plate of wastewater samples included PCR positive controls for each target assayed on the plate in 1 well, PCR negative no template controls in two wells, and extraction negative controls (consisting of water and lysis buffer) in two wells. PCR positive controls consisted of viral gRNA or gene blocks (Table S2). Results from replicate wells were merged for analysis. In order for a sample to be recorded as positive, it had to have at least 3 positive droplets.

Concentrations of RNA targets were converted to concentrations per dry weight of solids in units of copies (cp)/g dry weight using dimensional analysis. The total error is reported as standard deviations and includes the errors associated with the Poisson distribution and the variability among the 10 replicates. Three positive droplets across 10 merged wells corresponds to a concentration between ∼500-1000 cp/g; the range in values is a result of the range in the equivalent mass of dry solids added to the wells.

### Wastewater SARS-CoV-2 N gene

We obtained SARS-CoV-2 N gene concentrations at the POTW from a regional monitoring program^12^. These data were used in a supplementary manner to provide insight into the progress of the COVID-19 pandemic (see SM).

### Clinical data

California Sentinel Clinical Laboratories (hereafter, sentinel laboratories) test clinical specimens for influenza A (IAV) and influenza B (IBV), respiratory syncytial virus (RSV), parainfluenza viruses (HPIV) (1-4), human coronaviruses (HCoV) (229E, NL63, OC43, and HKU-1), human metapneumovirus (HMPV), and rhinovirus (HRV). Sentinel laboratories do not differentiate rhinovirus from other enteroviruses. Specimens tested by the laboratories may represent inpatient or outpatient samples of people receiving medical care. Positivity rates were calculated using data from all state sentinel laboratories and are reported by MMWR (Morbidity and Mortality Weekly Report) week and assigned to the first day of MMWR week. Positivity rates were also aggregated at the county-level for comparison (see SM).

### Statistical analysis

Kendall’s tau was used to test the null hypotheses (1) clinical specimen positivity rates are not associated with viral concentrations in wastewater solids, and (2) viral RNA concentrations were not associated with each other. Because case data are aggregated by MMWR week, we used median wastewater measurements from the same MMWR week to match clinical and wastewater data. To test correlations between wastewater viral RNA concentrations, wastewater data were smoothed by calculating a rolling three-point median (hereafter “smoothed wastewater concentrations”). A total of 50 Kendall’s tests of association were carried out so a p value of 0.001 was used to identify tau values significantly different from 0 based on Bonferroni’s correction. To compare viral RNA concentrations visually on a similar scale, we standardized their concentrations by subtracting the minimum and dividing by the range.

## Results

### QA/QC

Results are reported as suggested in the EMMI guidelines^19^. All extraction and PCR negative and positive controls performed as expected (negative and positive, respectively) with the exception of the positive control failure for an HCoV run including samples before 2/26/22; HCoV results prior to 2/26/22 were therefore omitted from the analysis. Median PMMoV concentrations across the samples were 1.6×10^9^ copies/g, similar to measurements previously reported for the plant^12^. In addition, PMMoV levels were stable across samples (interquartile range = 0.7×10^9^ cp/g) suggesting consistent fecal strength and RNA extraction efficiency. See SM for additional QA/QC details.

### Novel assay sensitivity and specificity

*In silico* analysis indicated no cross reactivity of the novel RT-PCR probe-based assays (Table 1) with sequences deposited in NCBI. The novel assays for HPIV, HMPV, HRV, and the HCoV were tested *in vitro* against panels of non-target viral gRNA as well as target gRNA (Table S2). No cross reactivity was observed.

**Table 1.**
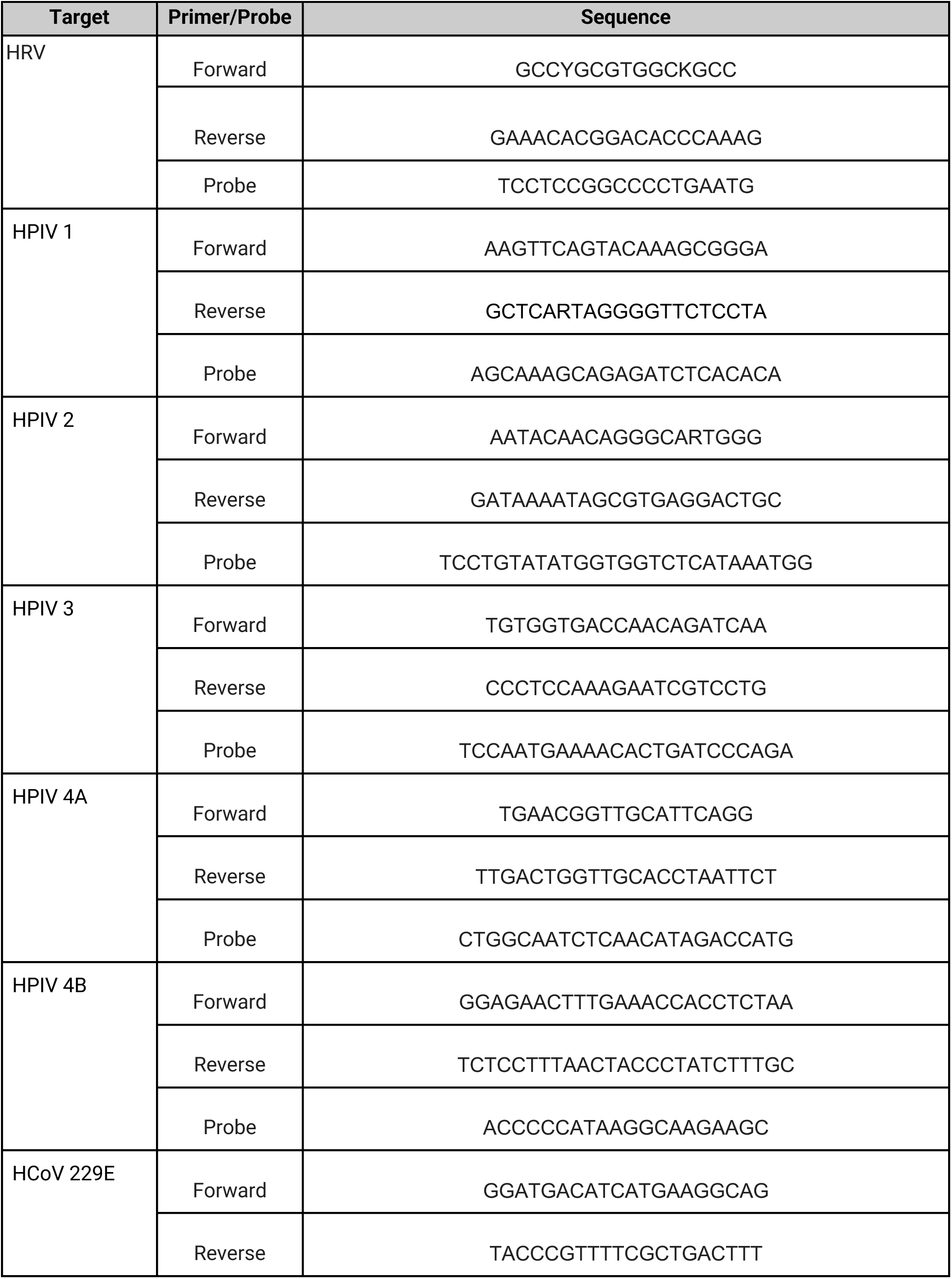
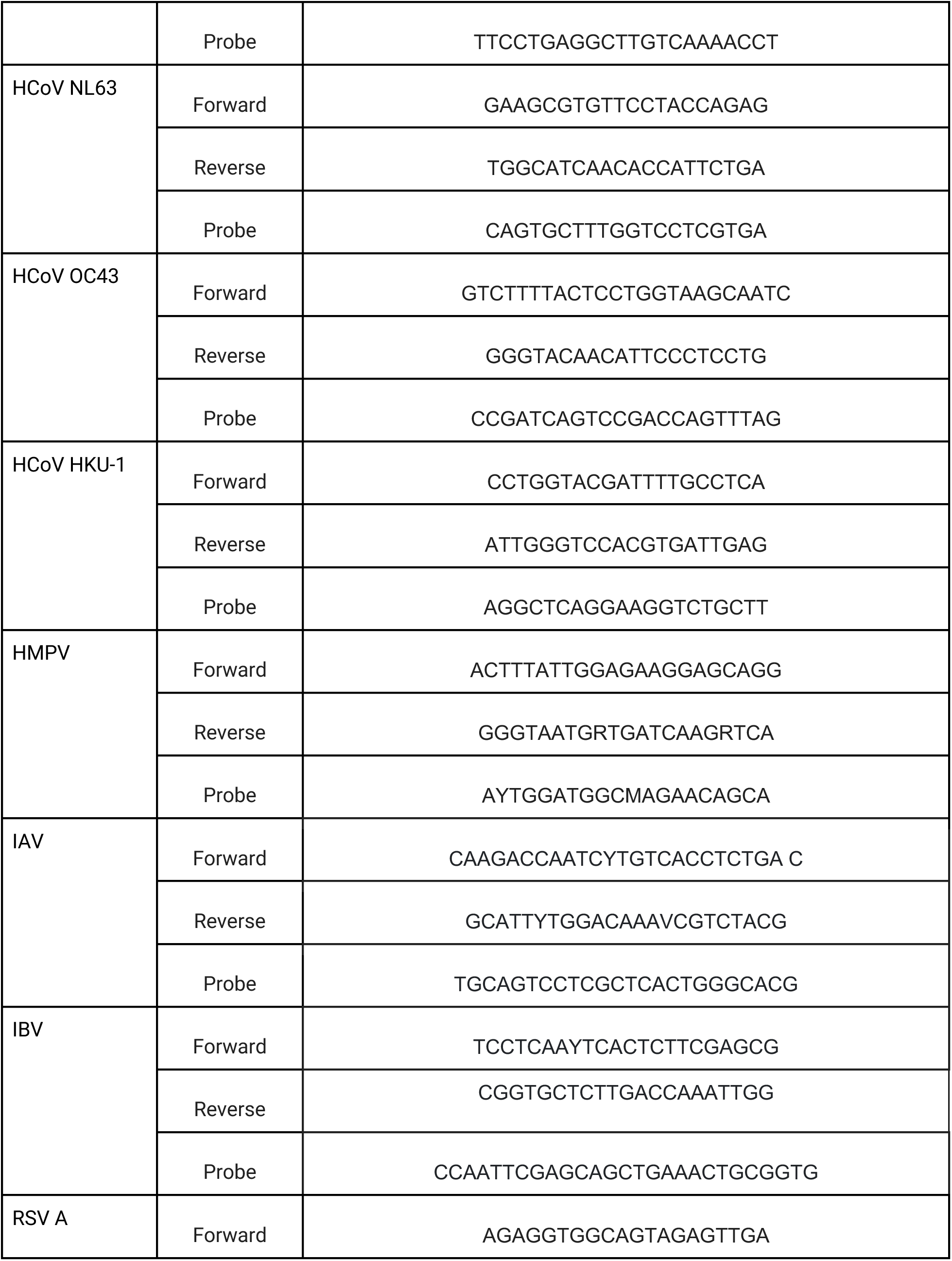
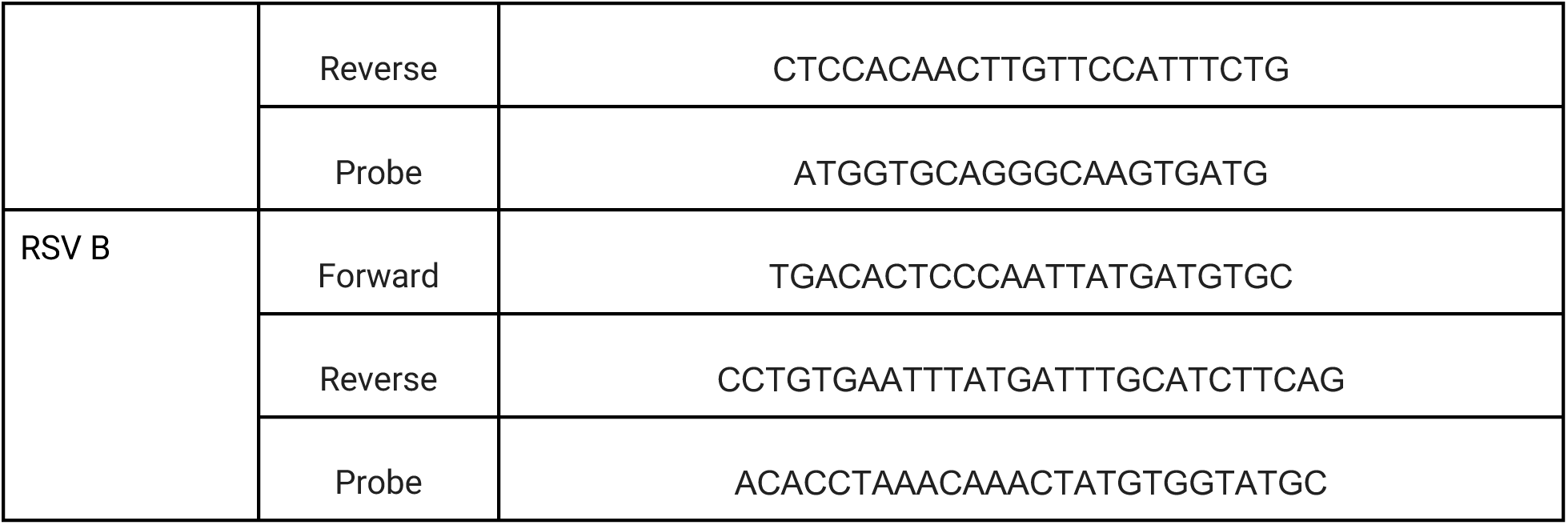
Primer and probe sequences for detection of respiratory viral RNA used in this study. Additional details of the target region of the genomes is the Supplementary Material. Primers and probes were purchased from IDT. All probes contained a fluorescent molecule and quenchers (5′ FAM and/or HEX/ZEN/3′ IBFQ). FAM, 6-fluorescein amidite; HEX, hexachloro-fluorescein; ZEN, a proprietary internal quencher from IDT; IBFQ, Iowa Black FQ. The Supplementary Material indicates whether HEX or FAM molecules were used.

### Respiratory virus RNA in wastewater solids

RNA from all viruses were detected in wastewater samples (Figure 1). Concentrations varied from non-detect to over 10^6^ copies/g. In order of highest to lowest, median concentrations were 35000 cp/g for HCoV, 8500 cp/g for HRV, 2500 cp/g for HPIV, 1700 cp/g for RSV B, 760 cp/g for HMPV, and non-detect for IAV, IAB, and RSV A (Table 2). While median IAV, IBV, and RSV A were non-detect, viral RNA from these viruses was detected in 28%, 6% and 14% of the samples, respectively. For context, median concentrations for SARS-CoV-2 was 48000 cp/g. In general, concentrations were lowest at the beginning of the study (February 2021), and increased until January 2022 at which time viral RNA for all viruses showed a steep drop in concentration. Coincidentally, January 2022 marked the onset of the Omicron BA.1 wave in the region which caused the highest COVID-19 reported incidence rates to date. After the steep drop off, viral RNA concentrations began to rise until the end of the time series with the exception of RSV B for which we did not observe a rebound. HCoV rebounded first when all other viral RNA concentrations, including those of SARS-CoV-2, were still at relatively low concentrations (Figure 2).

**Table 2.** Summary statistics of measurements of viral RNA in wastewater solids. Units of concentration is copies per gram dry weight of wastewater solids. RSV is respiratory syncytial virus, HCoV is the sum of all four non-SARS human coronaviruses (OC43, HKU-1, 229E, and NL63), HPIV is human parainfluenza viruses (1-4), HRV is human rhinovirus A, B and C, HMPV is human metapneumovirus, and IAV and IBV are influenza A and B viruses, respectively. SARS-CoV-2 data are included for context. # ND is number of samples for which the viral RNA was not detected. N is number of measurements made.

| Statistic | HPIV | HCoV | HMPV | HRV | RSV A | RSV B | IAV | IBV | SARS-CoV-2 |
| --- | --- | --- | --- | --- | --- | --- | --- | --- | --- |
| median | $3.5 \times 10^3$ | $3.5 \times 10^4$ | $7.6 \times 10^2$ | $8.5 \times 10^3$ | 0 | $1.7 \times 10^3$ | 0 | 0 | $4.8 \times 10^4$ |
| 25th %ile | $1.4 \times 10^3$ | $1.7 \times 10^4$ | 0 | $4.6 \times 10^3$ | 0 | 0 | 0 | 0 | $2.5 \times 10^4$ |
| 75th %ile | $6.3 \times 10^3$ | $5.6 \times 10^4$ | $2.0 \times 10^3$ | $1.7 \times 10^4$ | 0 | $5.4 \times 10^3$ | $6.6 \times 10^2$ | 0 | $1.3 \times 10^5$ |
| # of ND | 21 | 0 | 95 | 18 | 186 | 89 | 156 | 204 | 0 |
| N | 216 | 207 | 216 | 216 | 216 | 216 | 216 | 216 | 216 |

**Figure 1.**
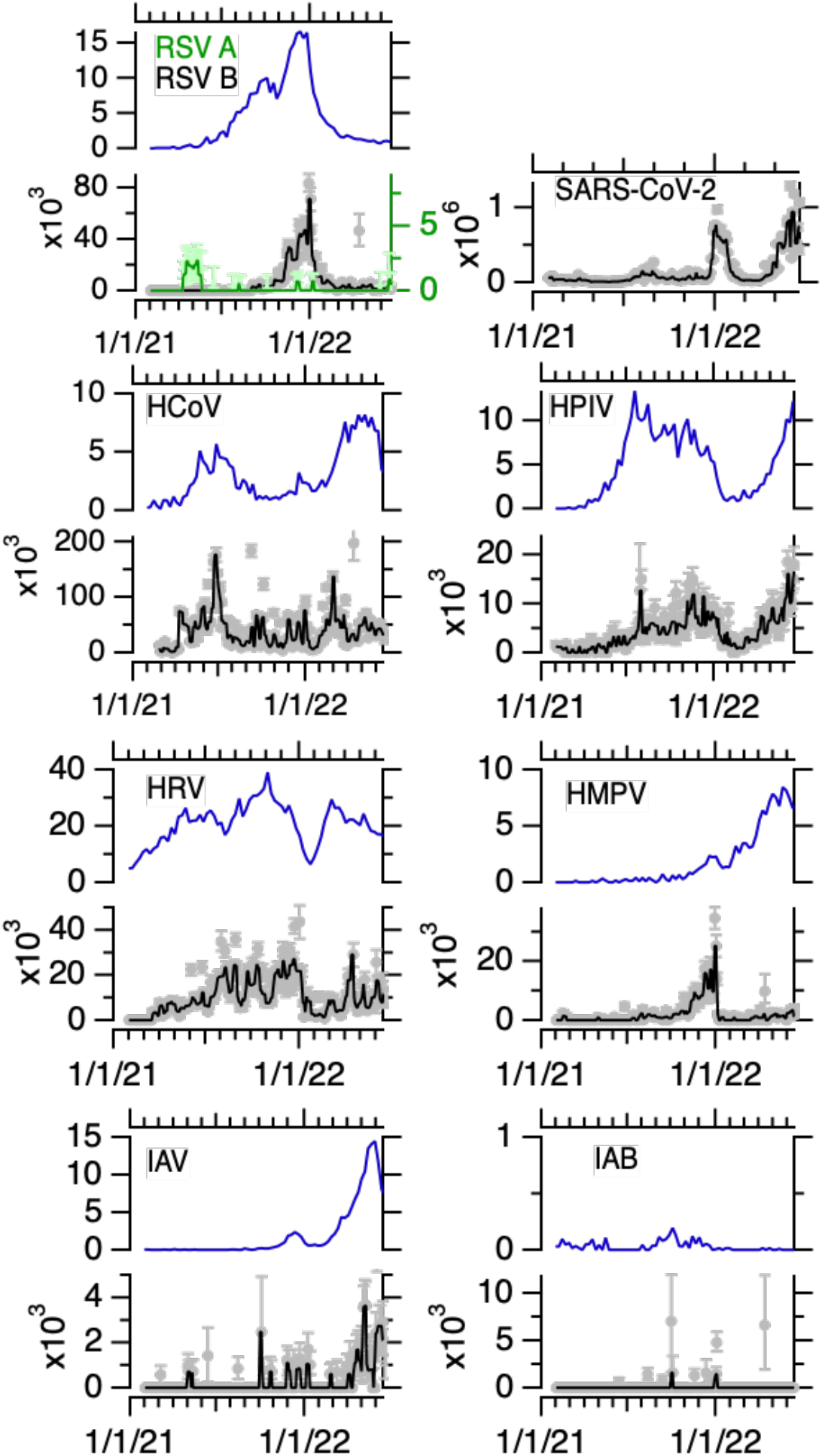
State-aggregated positivity rate reported as a percent from sentinel laboratories (top panel) and viral RNA in wastewater solids for each virus reported in copies per gram dry weight (bottom panel), exception is the top right panel which shows SARS-CoV-2 wastewater results for context. RSV is respiratory syncytial virus (A is shown in green and B in grey/black, RSV A scale is 1/10th that of RSV B and, like the RSV B axis, is scaled by 10^3^), HCoV is the sum of all four seasonal human coronaviruses (OC43, HKU-1, 229E, and NL63), HPIV is human parainfluenza viruses (1-4), HRV is human rhinovirus A, B and C, HMPV is human metapneumovirus, and IAV and IBV are influenza A and B viruses, respectively. Gray symbols represent measurements, error bars are 68% confidence intervals representing the total error around 10 replicates. The black line is the 3-point rolling median of the measurements. For HCoV there are three measurements located off scale (1.1×10^6^, 3.6×10^5^, and 5.1×10^5^ cp/g on 4/9/21, 6/24/21, and 3/3/22, respectively). For HPIV there is one measurement located off scale (3.6×10^4^ cp/g on 4/9/21). For HRV there is one measurement located off scale (3.6×10^5^ cp/g on 4/14/22). For IAV there are 3 measurements located off scale (2.1×10^4^, 7.5×10^4^, 5.1×10^5^ and 1.5×10^4^ cp/g on 9/30/21, 10/31/21, 3/3/22, and 4/14/22, respectively). Dates are in month/day/year format.

**Figure 2.**
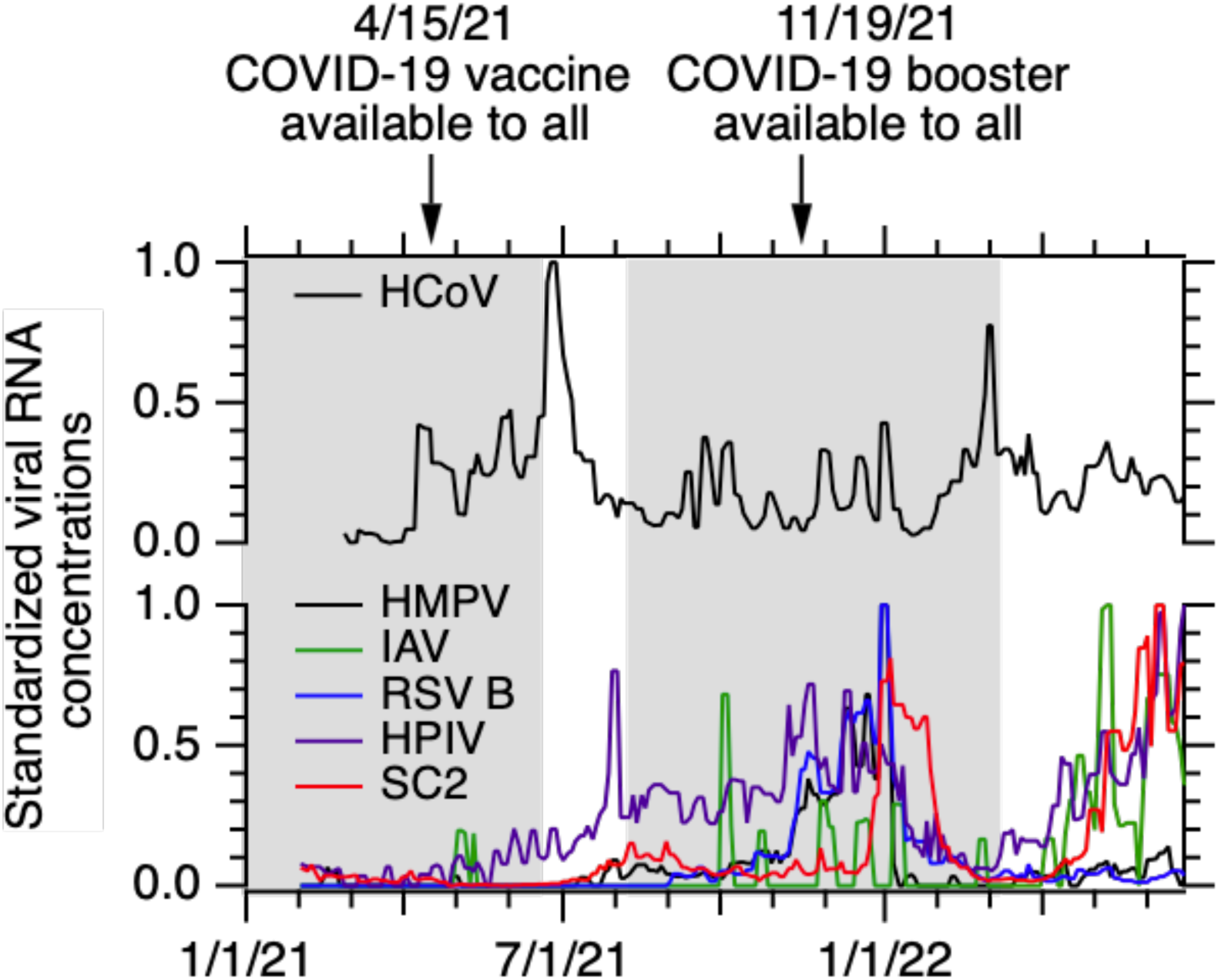
Standardized concentrations of viral RNA for all viruses except for RSV A and IBV which were rarely detected. Shaded background indicates periods of time that indoor masking mandates were in effect locally. Dates of COVID-19 vaccine availability are provided at top. Dates are in month/day/year format. Standardized concentrations were calculated by subtracting the minimum and dividing by the range. SC2 is SARS-CoV-2.

In order to assess which HCoV were circulating during the study, we measured concentrations of OC43, HKU-1, 229E, and NL63 RNA in nine samples spanning the time series (Figure 3). Nearly 100% of the total HCoV RNA was OC43 RNA in seven of the nine samples, all collected prior to April 2022. For the two samples collected thereafter, the contribution of OC43 RNA declined to 20-30% and contributions of 229E and HKU-1 increased from <10% to greater than 50% and 20-30%, respectively.

**Figure 3.**
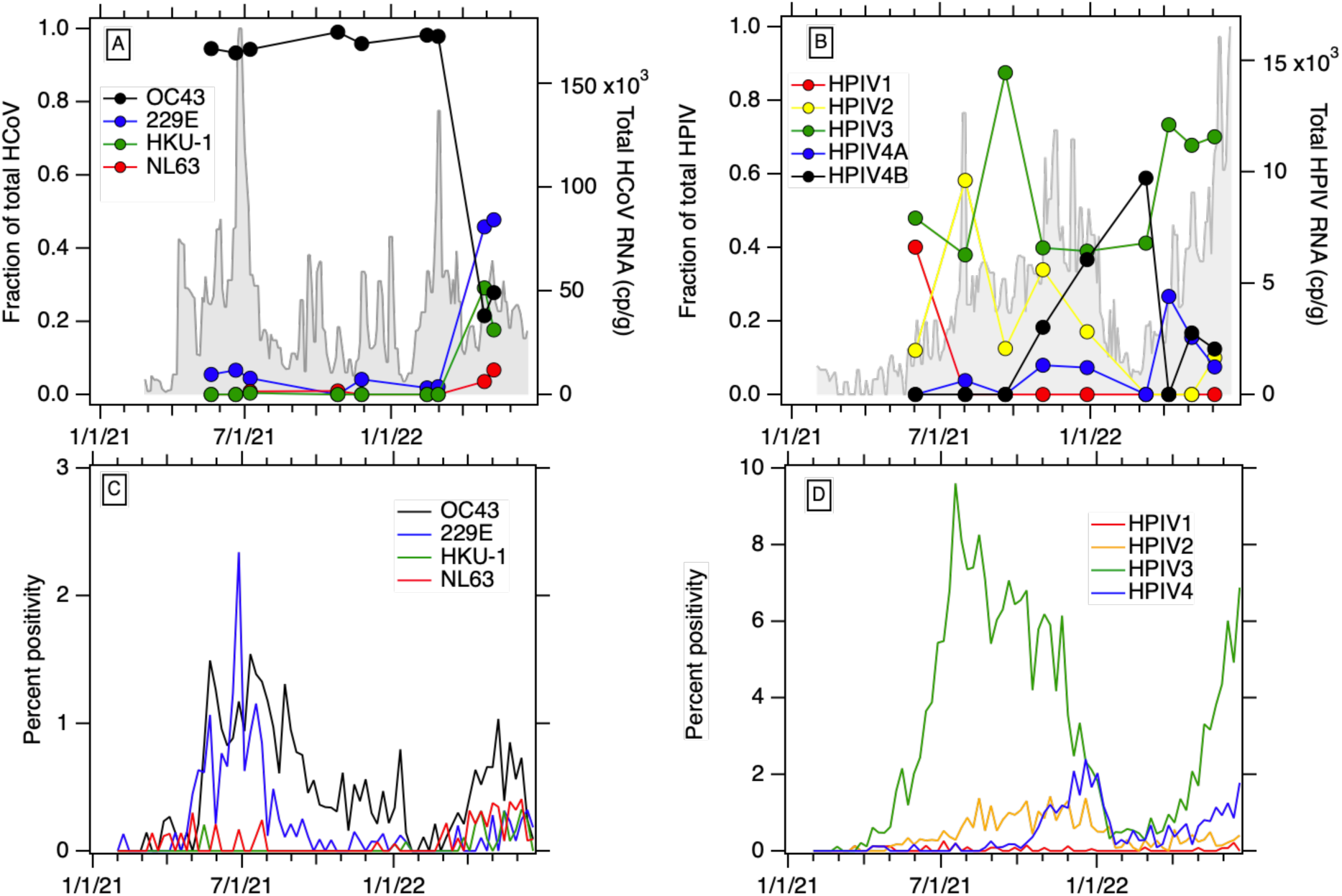
Data from wastewater solids on the (A) total concentration of HCoV RNA (shown as gray filled areas) and individual HCoVs (reported as fractions of total) and (B) total concentration of HPIV RNA (shown as gray filled areas) and individual HPIVs (reported as fractions of total). State-aggregated percent positivity for tests for (C) individual HCoVs and (D) individual HPIVs. Note that percent positivity for “unknown HCoV” and “unknown HPIV” from the sentinel surveillance system are not shown in panels C and D. Dates are in month/day/year format.

Similarly, we measured RNA concentrations of each HPIV (1, 2, 3, 4A, and 4B) in samples spanning the time series. RNA from all four HPIV were present; HPIV 3 dominated in most samples (40-90% of total HPIVs), although each HPIV 2 and 4B also dominated in one of the nine tested samples. HPIV 1 and 4A contributed the least to total HPIV present (median < 10%, Figure 3).

### Associations between viral RNA and confirmed case positivity rates

We tested whether state-aggregated positivity rates for respiratory viral infections was associated with the wastewater concentrations of viral RNA. Sentinel laboratories tested a median of 5918 (range 1809-14813) clinical specimens per week for influenza (IAV and IAB), 2419 (619-10907) for RSV, 1084 (574 - 1984) for HPIV, HMPV, and HRV, and 931 (469-1833) for HCoV. The associations between wastewater RNA concentrations and positivity rates were significant and positive for all viruses except for IBV. Kendall’s tau between positivity rates and wastewater concentration were 0.36 (p<10^−5^) for HRV, 0.45 (p<10^−5^) for IAV, 0.52 (p<10^−9^) for HPIV, 0.57 (p<10^−10^) for RSV (using RSV B wastewater concentrations as RSV A was mostly non-detect), 0.32 (p=0.00018) for HMPV, 0.32 (p=0.00011) for HCoV, and −0.010 (p=0.92) for IBV. Wastewater concentrations and positivity rates aggregated across viruses were positively correlated (tau = 0.50, p <10^−15^) (Figure S1). Results for county-aggregated positivity rates are reported in the SM (Figure S2).

State-aggregated positivity rates for individual HCoV infections are similar to observations of the relative occurrence of their RNA in wastewater solids (Figure 3). OC43 had the highest positivity relative to other HCoV, in agreement with the wastewater measurements. Wastewater indicates that 229E dominated at the end of the study (spring 2022), but this is not reflected in the clinical data.

Positivity rates for individual HPIV suggests HPIV 3 dominated HPIV infections throughout the study period; HPIV 2 and 4 also circulated with lower positivity rates. HPIV 1 was rarely detected in clinical specimens. This is in agreement with the wastewater data that indicated HPIV 3 dominated with HPIV 2 and 4B also present at relatively high levels, and HPIV 1 rarely detected (Figure 3).

### Associations between wastewater viral RNA concentrations

Viral RNA concentrations were associated with each other, as well SARS-CoV-2 N gene concentrations available from a regional monitoring program at the POTW (Table S4). Three-day smoothed wastewater concentrations of HPIV, HRV, HMPV, IAV, RSV B, and SARS-CoV-2 RNA were significantly correlated with each other (p<0.001) with tau from 0.18 (IAV and HRV) to 0.61 (HRV and HMPV). Given the low occurrence of IBV and RSV A RNA, it is not surprising that they were not correlated with other viral RNA concentrations. HCoV RNA concentrations were not correlated to concentrations of other viral RNA targets.

### Ethics

This study was reviewed by State of California Health and Human Services Agency Committee for the Protection of Human Subjects and determined to be Exempt.

### Role of funding source

CDC Foundation played no role in study design, execution, or result interpretation.

## Discussion

We detected RNA from all tested respiratory viruses in wastewater solids, including human rhinovirus, parainfluenza, metapneumovirus, influenza, respiratory syncytial virus, and seasonal human coronaviruses. Additionally, we detected four types of seasonal human coronaviruses including OC43, 229E, NL63, and HKU-1, and five different parainfluenza viruses (1, 2, 3, 4A, and 4B). Non-influenza respiratory viral RNA targets were measured using novel assays; these assays were found to be specific to the intended viral genomic targets when tested against a range of viruses. Previous studies have documented human rhinovirus^11,20^, RSV^13^, and influenza^14^ RNA in wastewater, but to our knowledge, no study to date has reported concentrations of other respiratory viruses.

RSV A RNA was rarely detected in wastewater, yet RSV B concentrations were >10,000 cp/g at times, and RSV B wastewater trends mirrored RSV clinical surveillance data. This suggests limited local co-circulation of the two RSV groups. Changes in dominant RSV strains have been documented to occur at continental^21^ and community scales^22^ with resultant changes in population immunity^23^. Sequencing of SARS-CoV-2 in wastewater has been successful at identifying variants^24^, and sequencing of RSV genomes from wastewater may yield additional insight into strain circulation and their potential susceptibility to different pharmaceutical interventions.

The association between respiratory viral RNA in wastewater solids and their respective virus clinical specimen positivity rates as available through a passive disease surveillance system suggests that wastewater data are representative of respiratory disease occurrence in the contributing population. This is despite limitations with passive disease surveillance, and the fact that data were aggregated across the entire state which obfuscates disease occurrence patterns at more local spatial scales. For example, state-aggregated HCoV case positivity suggests OC43 infections dominated HCoV infections, yet wastewater solids measurements suggest 229E dominated locally in spring of 2022. Similarly, based on wastewater measurements, HPIV 1 and 4 may have contributed to more HPIV infections locally than on average across the state.

There are uncertainties associated with understanding disease occurrence in a community through both clinical surveillance and wastewater testing. Clinical surveillance data is biased towards those receiving medical care and not all specimens are tested for all viruses, making it difficult to estimate disease occurrence from positivity rate data. Positivity rates may not be associated with disease occurrence in the same manner for all considered viruses. For wastewater, there is limited information on concentrations of viral RNA excreted from infected individuals into wastewater despite evidence showing that shedding does occur for viruses included in this study^25,26^. Given the current state of knowledge, it is difficult to know how inputs of viral RNA to the wastewater system differ between individuals infected with different viruses and with different disease severities.

Despite this, the association between viral RNA in wastewater solids and case positivity holds when data from the measured viruses are aggregated, suggesting that even across viruses,concentrations in wastewater solids are related to their different rates of disease occurrence. HCoV and HRV concentrations were highest across our study period, suggesting seasonal human coronavirus and human rhinovirus infections were most common. Conversely, IBV and RSV A concentrations were lowest suggesting limited circulation of those viruses. Future work is needed to document time varying patterns of respiratory viral RNA inputs from infected individuals to the wastewater system to enable modeling of the number of infected individuals in a sewershed from wastewater viral RNA concentrations^27^ and to enable modeling of epidemiological parameters like the reproductive number from wastewater data^28^.

Viral RNA concentrations in wastewater solids, including those of SARS-CoV-2, generally followed similar trends over time, with the exception of HCoV. Evident in the trends is a notable drop-off in concentrations of all viruses after the Omicron BA.1 surge in January 2022. The drop-off may suggest that disease mitigation measures practiced by the community in response to the Omicron surge (isolation due to illness, working remotely, wearing masks) reduced the spread of all respiratory viruses. Local indoor mask mandates ended shortly after the Omicon surge, and this change was followed by an increase in concentrations of all respiratory viruses in wastewater (Figure 3). HCoV, specifically OC43, was the first virus to re-appear in wastewater after the Omicon surge. Non-SARS-CoV-2 HCoV are highly transmissible with R0 values higher, on average, than influenza and rhinovirus^29^, but lower than RSV and similar to HPIV; there is limited R0 data for HMPV^15^. It is also possible that infection with Omicron reduced susceptibility to other respiratory viruses and contributed to the decrease in all viral activity; antibody cross immunity has been suggested to control temporal patterns of some respiratory viruses^5,30^.

Data generated from wastewater can be available within 24 hours of sample collection, and samples represent the entire contributing population - even those with asymptomatic infections - thereby overcoming various biases and the inherent reporting delays of the passive surveillance system. Wastewater surveillance for multiple respiratory viruses that commonly circulate can provide information at a community scale on the most likely causes of ARI. Although testing of clinical specimens is often not done for ARI, the viruses included in this panel have differing levels of likely severity and associated prevention measures, treatments, and complications. Therefore, more information about the abundances of circulating viruses can inform local clinical decision making including the prescription of drugs and testing, availability of community testing resources, and vaccination or behavior change campaigns. This information can also be communicated to the public, like a weather report, so the community and particularly vulnerable individuals can make informed decisions about behaviors that may put them at risk.

## Data Availability

Wastewater data are available publicly at the Stanford Digital Repository (https://doi.org/10.25740/vx726fw9373). State-aggregated surveillance data is available upon request from California Department of Health.

## Acknowledgements

This work is supported by a gift from the CDC Foundation. We acknowledge Payal Sarkar at RWF for overseeing sample collection. We acknowledge Erin Murray, Monica Sun, Tomás Leon, and Alexander Yu at California Department of Public Health for their assistance. This study was performed on the ancestral and unceded lands of the Muwekma Ohlone people. We pay our respects to them and their Elders, past and present, and are grateful for the opportunity to live and work here.

