## Supplemental Information for "Wastewater surveillance of human influenza, metapneumovirus, parainfluenza, respiratory syncytial virus (RSV), rhinovirus, and seasonal coronaviruses during the COVID-19 pandemic"

### Table of contents

**Supplementary Methods** 2

**Supplementary Results** 3

Table S1. Parameters used in Primer3Plus (https://primer3plus.com/). 5

Table S2. List of viruses included in this study, and the name of the gene target RT-PCR probe-based assays targeted. The list of non-target and target controls used to test assay specificity are provided. 6

Table S3. Samples chosen for analysis of different HCoVs and HPIVs. 8

Table S4. Kendall's test of association between smoothed wastewater solids concentrations of viral RNA. 8

Figure S1. Case positivity from clinical specimens submitted to sentinel laboratories aggregated across the state versus median concentration of viral RNA in wastewater solids for all viral targets for each MMWR week included in the study. 9

Figure S2. State-aggregated (blue) and county-aggregated (red) positivity rate reported as a percent from sentinel laboratories (top panel) and viral RNA in wastewater solids for each virus reported in copies per gram dry weight (bottom panel). 10

**References** 11

### Supplementary Methods

**Validation of newly designed respiratory virus probe-based RT-PCR assays.** Primers and probe sequences were screened for specificity *in silico* using NCBI Blast, and then tested *in vitro* against virus panels (NATtrol™ Respiratory Verification Panel NATRVP2-BIO or NATRVP2.1-BIO, Zeptomatrix, Buffalo, NY) that include chemically inactivated intact influenza viruses, parainfluenza viruses, adenovirus, rhinovirus, metapneumovirus, and coronaviruses, as well as various additional intact respiratory viruses, synthetic viral genomic RNA, or target cDNA sequences (Table S2). RNA was extracted from intact viruses using Chemagic Viral DNA/RNA 300 Kit H96 for Chemagic 360 (PerkinElmer, Waltham, MA). Nucleic acids were used undiluted as a template in digital droplet RT-PCR singleton assays in a single well. The concentration of nucleic acids used in the *in vitro* specificity testing was approximately 550 copies per reaction.

Immediately upon receipt at the lab, samples were centrifuged to dewater the solids as described elsewhere[1], and the dewatered solids were stored immediately at -80°C until analysis. The time of storage spanned from 4 to 60 weeks. Samples were removed from the freezer and thawed at 4°C overnight. Then, solids were suspended in DNA/RNA Shield (Zymo Research, Irvine, California, USA) at a concentration of 0.75 mg (wet weight)/ml. This concentration of solids was chosen as it was shown to alleviate inhibition in downstream RT-PCR[2]. An additional aliquot of dewatered solids was dried in an oven[1] to determine its dry weight so that measured concentrations of nucleic acid targets could be normalized to gram dry weight. RNA was extracted from 10 replicate aliquots of dewatered settled solids suspended in the DNA/RNA Shield using the Chemagic Viral DNA/RNA 300 kit H96 for the Perkin Elmer Chemagic 360 (Perkin Elmer, Waltham, MA) followed by PCR inhibitor removal with the Zymo OneStep-96 PCR Inhibitor Removal kit (Zymo Research, Irvine, CA) The pre-analytical methods described here are provided in detail in other publications and on protocols.io [1–5].

Nine samples spanning the duration of the time series were selected to measure each HCoV and HPIV, respectively (Table S3). Aliquots of extracted RNA were stored at -80°C for between 1 and 3 months before being thawed to measure concentrations of individual HCoVs and HPIVs.

Droplets were generated using the AutoDG Automated Droplet Generator (Bio-Rad, Hercules, CA). PCR was performed using Mastercycler Pro (Eppendforf, Enfield, CT) with with the following cycling conditions: reverse transcription at 50°C for 60 minutes, enzyme activation at 95°C for 5 minutes, 40 cycles of denaturation at 95°C for 30 seconds and annealing and extension at 59°C for 30 seconds, enzyme deactivation at 98°C for 10 minutes then an indefinite hold at 4°C. The ramp rate for temperature changes were set to 2°C/second and the final hold at 4°C was performed for a minimum of 30 minutes to allow the droplets to stabilize. Droplets were analyzed using the QX200 Droplet Reader (Bio-Rad). A well had to have over 10,000 droplets for inclusion in the analysis. All liquid transfers were performed using the Agilent Bravo (Agilent Technologies, Santa Clara, CA).

The ddRT-PCR methods applied to wastewater solids to measure PMMoV are provided in detail elsewhere[1] and are not repeated here. Here, we provide methods for the multiplexed respiratory virus assays which have not been reported previously. To obtain the total concentration of non-SARS-CoV-2 HCoV: the four HCoV assays were multiplexed with HCoV HKU1 and 229E in the FAM channel, and OC43 and NL63 in the HEX channel. To obtain the total concentration of HPIV the five HPIV assays were multiplexed: HPIV1, HPIV3, and HPIV 4A in the FAM channel, and HPIV 4B and HPIV 2 in the HEX channel. IAV (FAM), IBV (HEX), RSV A (FAM/HEX) were multiplexed using the triplex probe mixing approach. HMPV (FAM), HRV (HEX), and RSV B (FAM/HEX) were multiplexed using the triplex probe mixing approach. Finally, PMMoV (HEX) was measured in singleplex (ref). Using this approach, concentrations of individual genomic targets can be obtained from all assays except those for HCoV and HPIV - for those reactions, all positive droplets were combined to obtain concentrations of all HCoV and HPIV.

The nine samples chosen to measure each HCoV and HPIV individually were run as follows. 229E (FAM) and NL63 (HEX), and HKU-1 (FAM) and OC43 (HEX) were multiplexed. HPIV2 (FAM/HEX), HPIV3 (HEX), and HPIV 4A (FAM) were multiplexed , and HPIV1 (HEX) and HPIV 4B (FAM) were multiplexed.

Extracts from each of the 10 replicate nucleic acid extractions from each sample were run as template yielding 10 wells per sample. Each 96-well PCR plate of wastewater samples included PCR positive controls for each target assayed on the plate in 1 well, PCR negative no template controls in two wells, and extraction negative controls (consisting of water and lysis buffer) in two wells. PCR positive controls consisted of viral gRNA or gene blocks (Table S2).

ddRT-PCR was performed on 20 µl samples from a 22 µl reaction volume, prepared using 5.5 µl template, mixed with 5.5 µl of One-Step RT-ddPCR Advanced Kit for Probes (Bio-Rad 1863021), 2.2 µl of 200 U/µl Reverse Transcriptase, 1.1 µl of 300 mM dithiothreitol (DDT) and primers and probes mixtures at a final concentration of 900 nM and 250 nM respectively. Primer and probes for assays were purchased from Integrated DNA Technologies (IDT, San Diego, CA) (Table 1).

For the assay specificity testing, the respiratory virus assays were challenged against target and non-target (other respiratory viruses) viral gRNA (Table 1). Each PCR plate contained a negative PCR NTC.

Thresholding was done using QuantaSoft™ Analysis Pro Software (Bio-Rad, version 1.0.596). In order for a sample to be recorded as positive, it had to have at least 3 positive droplets.

**Regional monitoring program SARS-CoV-2 N gene and PMMoV gene concentrations.** The wastewater solids samples from RWF used in this study were collected as part of a larger wastewater monitoring program focused on measuring SARS-CoV-2 RNA in daily samples that were not stored prior to analysis. Detailed methods for that program have been published[1] and some of those results from samples included in this study have been published[1–5]. Herein, we include data on SARS-CoV-2 N gene concentrations in a supplementary manner to provide insight into the progress of the COVID-19 pandemic during this study, and data on PMMoV to assess how storage of the samples potentially affected viral RNA quantification.

### Supplementary Results

**QA/QC for wastewater samples.** Each plate run included negative extraction and RT-PCR controls, and positive extraction and PCR controls. In order for a plate to pass QA/QC, the number of positive droplets across merged negative extraction and PCR well could be no larger than 2 for any plate except the PMMoV plate where we allowed up to 13 positive droplets. The larger number of droplets in negative controls were allowed because PMMoV is a high copy number target with typically 71,000 (median across all samples) positive droplets for the samples; therefore addition of up to 13 positive droplets would change the percent of positive droplets by less than 0.02% and have minimal effect on the measured concentration. The number of positive droplets for the positive controls had to be greater than 3 droplets yielding a positive result. All plates passed this threshold. Median PMMoV concentrations across the samples were $1.6 \times 10^9$ copies/g similar to measurements previously reported for the plant[1]. In addition, levels were stable across all samples (IQR = $0.7 \times 10^9$ cp/g) suggesting consistent fecal strength and RNA extraction efficiency. The concentration of PMMoV obtained in this study was compared to the concentration of PMMoV measured in the same samples from fresh wastewater solids from our routine monitoring program. The median ratio of these concentrations was 1.1 (interquartile range IQR = 0.7). This suggests that the storage of the solids in this study at -80°C and their subsequent freeze thaw had limited impact on PMMoV RNA concentrations. We therefore assume that the impact was similarly minimal for the other targets. Results for HCoV for samples collected prior to 2/26/22 (month/day/year) were omitted from the analysis owing to failed positive controls.

**Summary of Santa Clara County sentinel lab data (county-aggregated sentinel lab data) and its association with wastewater measurements.** The wastewater treatment plant serves approximately 75% of the population of Santa Clara County, suggesting that disease incidence data from sentinel laboratories in the county would be a good proxy for disease incidence in the sewershed. However, the county-aggregated surveillance data was sparse, particularly for non-SARS HCoV, for which no clinical specimens were assayed. We therefore opted not to present the results from these analysis in the main paper, but include them herein for completeness.

Sentinel laboratories in Santa Clara county tested a median of 20 (range 4-935) specimens per MMWR week for influenza (IAV and IAB) and RSV, and a median of 7 (0 - 634) for HPIV, HMPV, and HRV. No clinical specimen was tested for HCoV. Wastewater viral RNA concentrations were positively associated with the positivity rates for the virus in clinical specimens processed by the county sentinel laboratories for IAV, HPIV, and RSV. Kendall's tau between positivity rates and median wastewater concentrations were 0.54 ($p=9x10^{-8}$) for IAV, 0.44 ($p=10^{-6}$) for HPIV, and 0.74 ($p=2x10^{-16}$) for RSV. Clinical specimen testing does not distinguish between RSV A and B, so wastewater RSV B concentration was used as a variable in the test of association as RSV A were mostly non-detect (Table 3). There was no reported positive clinical specimen for IBV and HCoV so we could not perform a test of association. There was not a correlation between HMPV RNA concentrations in wastewater and HMPV case positivity (tau = 0.67, p = 0.5) or HRV RNA wastewater concentrations and HRV case positivity (tau = 0.23, p=0.005).

| **Parameter** | **Values** |
|---|---|
| Primer size | min 15, optimum 20, max 36 |
| Primer melting temperature | min 50°C, optimal 60°C, max 65°C |
| GC% content: | min 40%, optimal 50%, high 60% |
| concentration of divalent cations | 3.8 mM |
| concentration of dNTPs | 0.8 mM |
| Internal oligo size | size min 15, optimal 20, max 30 |
| Internal oligo: Melting temp | min 62°C, optimal 63°C, max 70°C |
| Internal Oligo: GC% | min 30%, optimal 50%, max 80% |

**Table S1.** Parameters used in Primer3Plus (https://primer3plus.com/).

| **Virus(es) (abbreviation used in the paper** | **Genomic target** | **Non-target testing (negatives)** | **Target testing (positives)** |
|---|---|---|---|
| Pan Rhinovirus A/B/C (HRV) | Untranscribed region | NATRVP2-BIO | Clinical samples sequenced and classified as HRV A, B and C |
| Human Parainfluenza (HPIV) 1 | Hemagglutinin-neuraminidase (HN) gene | NATRVP2-BIO | Intact HPIV 1 virus (ATCC VR-94) |
| HPIV 2 | HN gene | NATRVP2-BIO | Intact HPIV 2 virus (ATCC VR-92) |
| HPIV 3 | Fusion protein (F) gene | NATRVP2-BIO | Intact HPIV 3 virus (ATCC VR-93) |
| HPIV 4A | Untranscribed region | NATRVP2-BIO | Intact HPIV 4A virus (ATCC VR-1378) |
| HPIV 4B | HN gene | NATRVP2-BIO | Intact HPIA 4B virus (ATCC VR-1377) |
| Human Coronavirus (HCoV) 229E | Nucleoprotein (N) gene | NATRVP2-BIO | Synthetic HCoV 229E gRNA (TWIST 103011) |
| HCoV NL63 | N gene | NATRVP2-BIO | Synthetic HCoV NL63 gRNA (TWIST 103012) |
| HCoV OC43 | N gene | NATRVP2-BIO | Synthetic HCoV OC43 RNA (TWIST 103013) |
| HCoV HKU-1 | N gene | NATRVP2-BIO | IDT gene Block |
| Human Metapneumovirus (HMPV) | RNA directed RNA polymerase (L) gene | NATRVP2.1-BIO | Synthetic HMPV RNA (ATCC VR-3250SD) and intact HMPV virus (Zepto NATRVP-IDI) |
| Influenza B (IBV) | Membrane (M1) gene | NATRVP2.1-BIO | Synthetic IBV gRNA (TWIST 103003) and intact IBV virus (Zepto NATFVP-NNS) |
| RSV A | N gene | NATRVP2.1-BIO | Intact RSV A virus (Zepto NATFVP-NNS) |
| RSV B | N gene | NATRVP2.1-BIO | Intact RSV B virus (Zepto NATFVP-NNS) |

**Table S2.** List of viruses included in this study, and the name of the gene target RT-PCR probe-based assays targeted. The list of non-target and target controls used to test assay specificity are provided. All non-target controls are virus panels sold by Zeptomatrix ("Zepto", Buffalo, NY). The vendor Twist Biosciences ("Twist") is located in South San Francisco, CA. ATCC is American Type Culture Collection.

| Virus group | Samples chosen for analysis of all members of virus group |
|---|---|
| HCoV | 5/20/21, 6/20/21, 7/8/21, 10/26/21, 11/28/21, 2/15/22, 3/1/22, 4/28/22, 5/10/22 |
| HPIV | 6/1/21, 8/1/21, 9/19/21, 11/4/21, 12/28/21, 3/10/22, 4/7/22, 5/5/22, 6/2/22 |

**Table S3.** Samples chosen for analysis of different HCoVs and HPIVs. Dates are provided in month/day/year format.

| | HMPV | IAV | IBV | HRV | RSV A | RSV B | HPIV | HCoV |
|---|---|---|---|---|---|---|---|---|
| IAV | **0.23** **$p<10^{-4}$** | | | | | | | |
| IBV | 0.15 $p=0.01$ | 0.074 $p=0.3$ | | | | | | |
| HRV | **0.51** **$p<10^{-15}$** | **0.18** **$p=0.0009$** | 0.15 $p=0.008$ | | | | | |
| RSV A | -0.15 $p=0.009$ | 0.059 $p=0.3$ | -0.055 $p=0.40$ | -0.06 $p=0.30$ | | | | |
| RSV B | **0.48** **$p<10^{-15}$** | **0.23** **$p<10^{-4}$** | 0.17 $p=0.004$ | **0.27** **$p<10^{-7}$** | -0.13 $p=0.02$ | | | |
| HPIV | **0.61** **$p<10^{-15}$** | **0.38** **$p<10^{-12}$** | 0.12 $p=0.04$ | **0.54** **$p<10^{-15}$** | -0.13 $p=0.02$ | **0.43** **$p<10^{-15}$** | | |
| HCoV | -0.10 $p=0.04$ | 0.10 $p=0.06$ | 0.15 $p=0.007$ | -0.0046 $p=0.9$ | 0.054 $p=0.33$ | -0.076 $p=0.1$ | -0.045 $p=0.3$ | |
| SARS-CoV-2 | **0.35** **$p<10^{-12}$** | **0.33** **$p<10^{-9}$** | 0.12 $p=0.04$ | **0.24** **$p<10^{-6}$** | -0.15 $p=0.006$ | **0.37** **$p<10^{-13}$** | **0.47** **$p<10^{-15}$** | -0.13 $p=0.004$ |

**Table S4.** Kendall's test of association between smoothed wastewater solids concentrations of viral RNA. Tau and the associated p value are provided. To account for multiple hypothesis testing (50 tests were completed), a p value cut off of 0.001 (based on Bonferroni's correction) was used to identify correlations statistically different from 0.

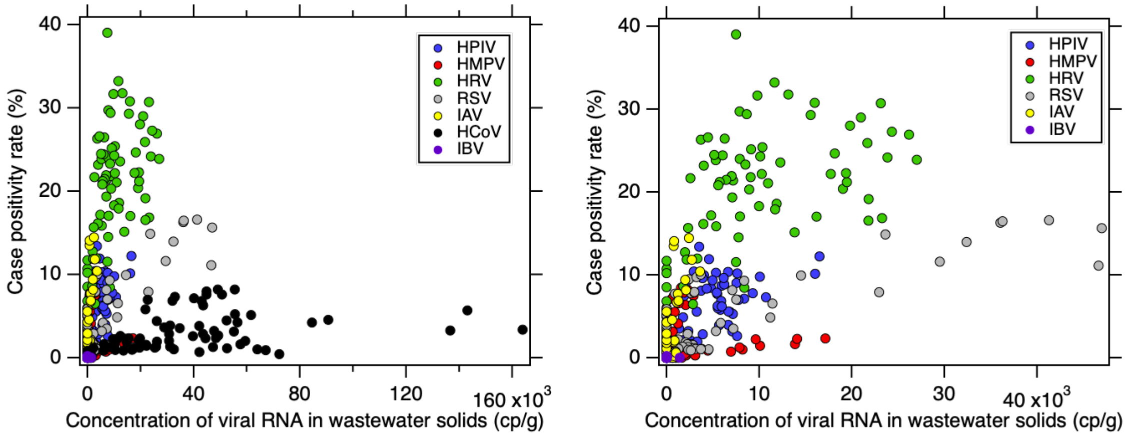


**Figure S1.** Case positivity from clinical specimens submitted to sentinel laboratories aggregated across the state versus median concentration of viral RNA in wastewater solids for all viral targets for each MMWR week included in the study. RSV B was used to represent RSV in wastewater solids. Left panel is all viruses, right panel excludes seasonal HCoV data to allow visualization of other data more easily.

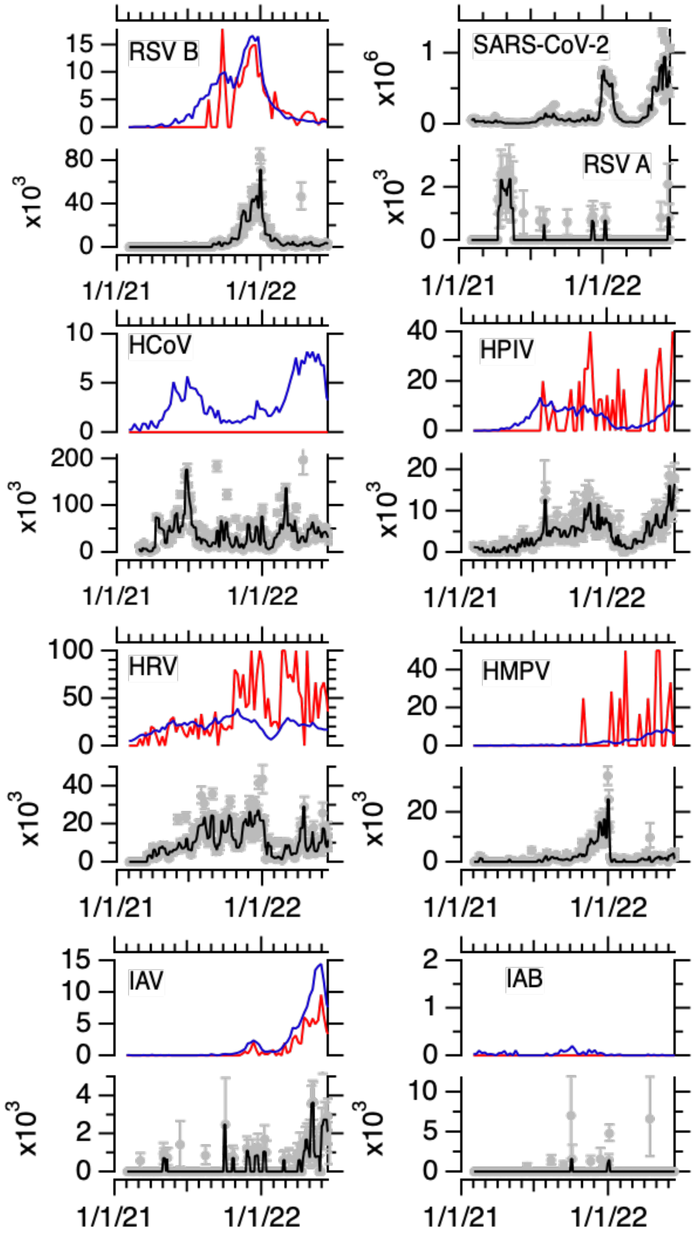


**Figure S2.** State-aggregated (blue) and county-aggregated (red) positivity rate reported as a percent from sentinel laboratories (top panel) and viral RNA in wastewater solids for each virus reported in copies per gram dry weight (bottom panel), exception is the top right panel which shows RSV A and SARS-CoV-2. RSV is respiratory syncytial virus, HCoV is the sum of all four non-SARS human coronaviruses (OC43, HKU-1, 229E, and NL63), HPIV is human parainfluenza viruses (1-4), HRV is human rhinovirus A, B and C, HMPV is human metapneumovirus, and IAV and IBV are influenza A and B viruses, respectively. SARS-CoV-2 is shown for context. Gray symbols represent measurements, error bars are 68% confidence intervals representing the total error around 10 replicates. The black line is the 3-point rolling median of the measurements. For HCoV there are three measurements located off scale ($1.1 \times 10^6$, $3.6 \times 10^5$, and $5.1 \times 10^5$ cp/g on 4/9/21, 6/24/21, and 3/3/22, respectively). For HPIV there is one measurement located off scale ($3.6 \times 10^4$ cp/g on 4/9/21). For HRV there is one measurement located off scale ($3.6 \times 10^5$ cp/g on 4/14/22). For IAV there are 3 measurements located off scale ($2.1 \times 10^4$, $7.5 \times 10^4$, $5.1 \times 10^5$ and $1.5 \times 10^4$ cp/g on 9/30/21, 10/31/21, 3/3/22, and 4/14/22, respectively). Dates are in month/day/year format.
